# Trends of COVID-19 pediatric admissions number during the first 24 weeks of COVID-19 vaccination in Rio de Janeiro, Brazil

**DOI:** 10.1101/2021.09.09.21263140

**Authors:** André Ricardo Araujo da Silva, Mônica del Monaco Esteves, Bernardo Rodrigues Rosa de Carvalho, Cristina Vieira Souza, Cristiane Henriques Teixeira

## Abstract

**Objective:** To describe trends of COVID-19 pediatric admissions number during the first 24 weeks of COVID-19 vaccination.

**Design:** A retrospective study was conducted in children (0-18 years), admitted in two pediatric hospitals of Rio de Janeiro city, between January 17 and July 3, 2021 with confirmed COVID-19 by reverse transcription polymerase chain reaction or serological tests. Trends of COVID-19 pediatric admissions number during the first 24 weeks of COVID-19 vaccination in Rio de Janeiro, Brazil and the pre-vaccine period were measured by linear regression.

**Participants:** Children admitted in pediatric hospitals in Rio de Janeiro, city, Brazil

**Results:** The number of total admitted patients (with all diseases) were 5340 during the pre-vaccine period, being 94 (1.8%) of them with confirmed COVID-19, and 4182 children admitted during the vaccine period, with 86 confirmed COVID-19 patients (2.1 %) (p=0.29). Media of cases admitted per/week were 2.02 in pre-vaccine period and 3.6 during the first 24 weeks of COVID vaccination (p=0.009). One death was reported in the pre-vaccine period and four in the vaccine period (p=0.14). Trends of increase in the number of admitted cases were verified both in the pre-vaccine period as in the vaccine period, being more expressive in the last one.

**Conclusion:** There was trend of increase in number of children admitted with confirmed COVID-19 during the first 24 weeks of COVID-vaccination in Rio de Janeiro, city. Considering that few people were fully vaccinated, reducing of number of admitted children with confirmed COVID-19 was not verified.

## Introduction

Starting in December 2020 in specific countries as United Kingdom and USA, COVID vaccination is one of the most promising strategies to control COVID-19 pandemics associated to non-pharmaceutical interventions (NPIs). ^1,2^

Despite children were less affected by COVID-19, severes cases of respiratory syndrome distress and multisystem inflammatory syndrome (MIS-C) are described, causing hospitalization and deaths. ^3,4^

Vaccines against SARS-COV-2 are available just for children above 12 years old age and initial analysis of a systematic vaccination progra ms verified reduction on hospital admissions and mortality in older adults. ^5,6^ In Brazil and specifically in Rio de Janeiro state, COVID-19 vaccination has started in January 19, 2021 (Brazilian epidemiological week 3) and we speculate an indirect effect of adult vaccination contributing to modify the trend of number of admitted children due to confirmed COVID-19 in pediatric hospitals. Considering these aspects, our aim is to describe COVID-19 pediatric admission number, during the first 24 weeks of COVID-19 vaccination.

## Methods

### Study design and setting

We conducted a retrospective cohort study in two pediatric hospitals in Rio de Janeiro, city, Brazil. Both hospitals are private units destined exclusively to pediatric patients.

Unit 1 is a 135-bed hospital located at North zone of the city, that receives clinical and surgical patients referred from its own emergency and from other services. Unit 2 is a 39-bed hospital, located at South zone of the city, with the same profile of unit 1.

### Period of the study

Two periods were analysed: the first one comprised the first day of Brazilian epidemiological week 13 (March 22, 2020) and the last day of epidemiological week 2 of 2021 (January 2, 2021). This period corresponds to the period without any COVID vaccine available in the city. The second period comprises the first day of epidemiological week 3 of 2021 (January 17^th^) and last day of epidemiological week 26 of the same year (July 3^rd^). This period corresponds to the first 24 weeks after beginning of vaccination in Rio de Janeiro city.

In both periods studied, higher number of COVID-19 cases were detected in the city. ^7^

### Inclusion and exclusion criteria

All patients admitted with COVID-19 in both hospitals were included in analysis. Children admitted but transferred to the other hospitals within the first 24h were excluded.

### COVID-19 vaccines available and priority groups

Four vaccines were available during the first 24 weeks by emergency use in Brazil: ChAdOx1 nCoV-19, BNT162b2, Ad26.COV2.S and CoronaVac. ^6^ The first doses of vaccines were administered in Rio de Janeiro, in January 20, 2021, first doses of Ad26.COV2.S were administered in the city, after June 26, 2021.

Until July 3, 2021, the following groups received at least one dose of vaccine: persons older than 43 years and healthcare workers (all ages) with direct contact with patients. No vaccine was available for children.

### Diagnostic of COVID-19 infection

A patient was considered as a confirmed case of COVID-19 in the presence of positive polymerase chain reaction (PCR) or positive antigen. Positive IgM or IgG serology were considered as evidence of infection in patients with multisystem inflammatory syndrome. For epidemiological analyses, we considered the date of beginning of the symptoms.

### Data source and statistical analysis

The two periods were analysed regarding the following variables: Total number of admitted patients (with all diseases), total number of admitted patients with confirmed COVID-19 and number of deaths

A descriptive analysis was performed using Microsoft Excel. When appropriate, Z test was performed to compare proportions, t test for compare means and Mann– Whitney U test for continuous variables. A value of p less than .05 were considered as statistical significant. Trends of COVID admissions were measured using linear regression, considering the period studied.

### Ethical aspects

The study was submitted and approved by Ethics Committee of Faculty of Medicine (Universidade Federal Fluminense) and Prontobaby Group, under number 4.100.232 dated from June 20, 2020. A consent statement for the use of data from the patients or their parents or guardians were required for each children included.

## Results

Until July 2, 2021, 9522 children were admitted in both hospitals studied. In the table 1 we show number of hospitalized children in the period pre and during COVID vaccination, number of confirmed COVID-19 cases and number of deaths.

**Table 1.** Total number of admissions, number of confirmed COVID-19 cases and deaths in pre and during vaccination period in two pediatric hospitals (Rio de Janeiro, Brazil)

|  | Pre-vaccine period | During<br>vaccine period | Total | P value |
| --- | --- | --- | --- | --- |
| N° of admitted children | 5340 | 4182 | 9522 |  |
| N° of confirmed cases (%) | 94 ( 1.8) | 86 ( 2.1) | 180 (1.9) | 0.29 |
| N° of deaths | 1 | 4 | 5 | 0.14 |

The first case of confirmed COVID was admitted within the Brazilian epidemiological week 13, in a children with fever and dyspnoea. In all the five deaths, children had a previous comorbidity (obesity and leukemia, 2 cases each one and neurological chronic disease).

The trends of number of admitted children with confirmed COVID is showed in graph 1. Media of admitted cases/week was 2.2 in the pre-vaccine period and 3.6 during the vaccine period.

**Graph 1.**
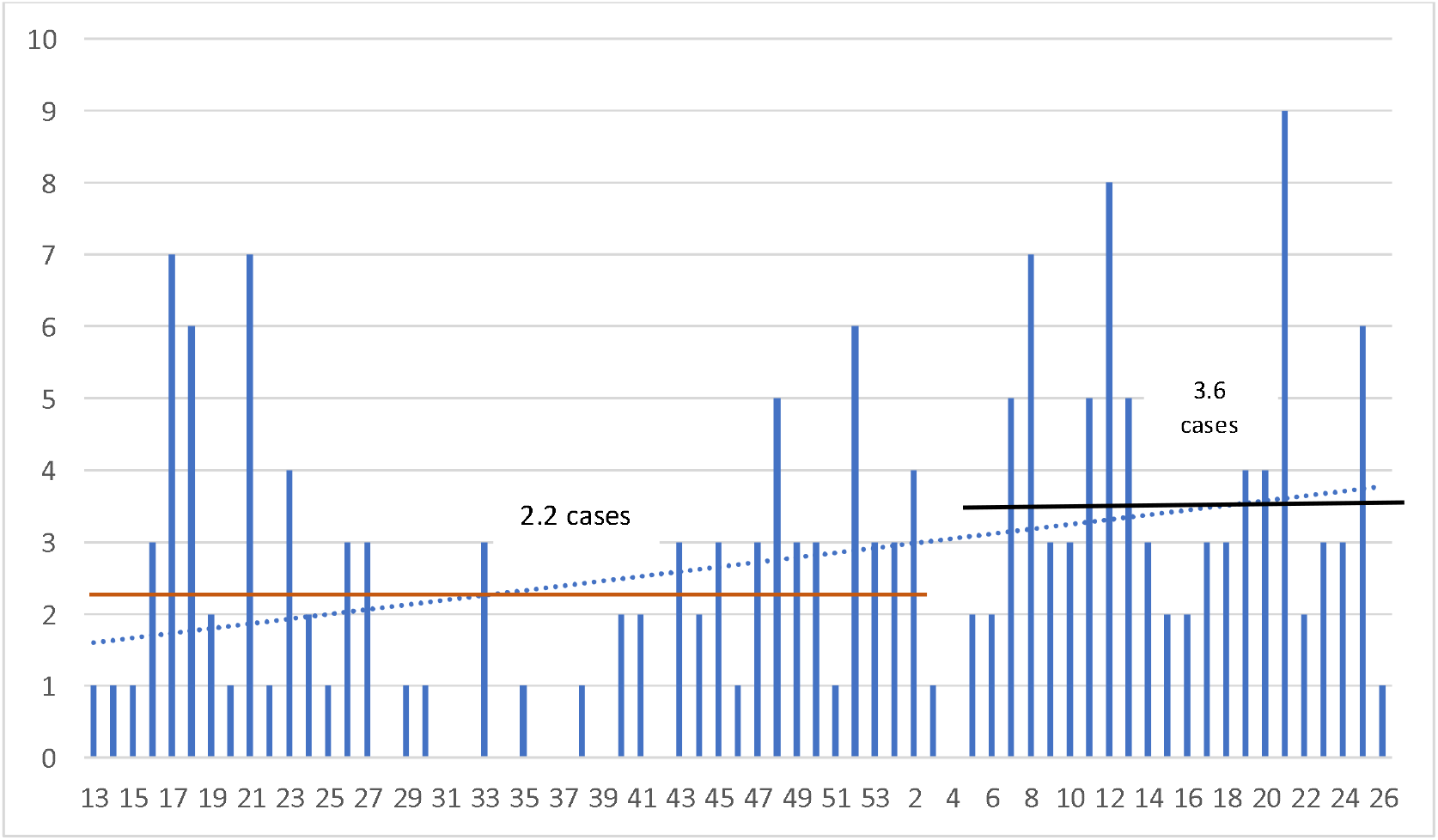
Trends of number of admitted children in two pediatric hospitals, according to the Brazilian epidemiological week (2020-2021) Blue continuous columns- number of cases/epidemiological week Blue traced-line- Linear trend

## Discussion

Until the end of August 2021, the COVID pandemics stills in course, causing more than 218 million of cases and 4,5 million of deaths around the world. ^8^ Considering that non-pharmaceutical interventions mitigate the spread of disease, effective vaccines are necessary to interrupt the pandemics. ^2,9^

In countries like Israel and United Kingdom, in which a large percentage of adult were fully vaccinated, it was verified effectiveness against SARS-CoV-2 asymptomatic infection, symptomatic infection, and COVID-19-related hospitalisation, severe or critical hospitalisation, and death for the adult population and specific groups as healthcare-workers. ^6,10^

In our study we proposed to verify trends of number of admitted children due to COVID-19, speculating an indirect positive effects of COVID-19 in Rio de Janeiro during the first 24 weeks of vaccination in the city, even considering that vaccine was not available for children.

We verified that relative percentage of admitted children with confirmed COVID-19 compared with total of admission was statistically similar in pre and during vaccination period as well number of deaths. This finding corroborate that COVID-19 is a mild/moderate disease in children, causing few deaths, mainly in patients with previous disease. ^3,4^

When trends of number of admitted children with confirmed children were analysed, curiously we verified more cases admitted/week during the period vaccination compared with the pre-vaccine period. Three possible reasons could explain these results: the first one is the introduction and circulation of Delta variant in the city, causing more cases including children (painel Rio de janeiro), the second one is available of more tests for COVID-19 detection in the city, mainly antigen-based tests and the last one is the few amount of population received fully vaccination.

Our research was conducted in two reference pediatric hospitals of Rio de Janeiro which could be considered a limitation of the study, but considering that hospitals received patients from all neighborhood of the city, the results could reflect the true dynamics of virus circulation in the city. The second limitation is disponibility of different vaccines, with different rates of effectiveness and unknown effects for long-term protection against Delta variant, which was the most one prevalent in Rio de Janeiro at the moment of the current research.

## Conclusion

We verified trends of increase in number of children admitted with confirmed COVID-19 during the first 24 weeks of COVID-vaccination in Rio de Janeiro, city. Considering that few people were fully vaccinated, reducing of number of admitted children with confirmed COVID-19 was not not verified.

## Data Availability

All data will be available under request

## Acknowlegdment

We thank to Mario Eduardo Viana and Luisa Benigno Barbosa Araujo da Silva for supporting this research.

## References

1) Ramos AM, Vela-Pérez M, Ferrández MR, Kubik AB, Ivorra B. Modeling the impact of SARS-CoV-2 variants and vaccines on the spread of COVID-19. Commun Nonlinear Sci Numer Simul. 2021 Nov;102:105937. doi: 10.1016/j.cnsns.2021.105937. Epub 2021 Jun 24. PMID: 34188432; PMCID: PMC8223013.

2) Haug N, Geyrhofer L, Londei A, Dervic E, Desvars-Larrive A, Loreto V, Pinior B, Thurner S, Klimek P. Ranking the effectiveness of worldwide COVID-19 government interventions. Nat Hum Behav. 2020 Dec;4(12):1303–1312. doi: 10.1038/s41562-020-01009-0. Epub 2020 Nov 16. PMID: 33199859.

3) Mantovani A, Rinaldi E, Zusi C, Beatrice G, Saccomani MD, Dalbeni A. Coronavirus disease 2019 (COVID-19) in children and/or adolescents: a meta-analysis. Pediatr Res. 2021 Mar;89(4):733–737. doi: 10.1038/s41390-020-1015-2. Epub 2020 Jun 17. PMID: 32555539.

4) Kaushik A, Gupta S, Sood M, Sharma S, Verma S. A Systematic Review of Multisystem Inflammatory Syndrome in Children Associated With SARS-CoV-2 Infection. Pediatr Infect Dis J. 2020 Nov;39(11):e340–e346. doi: 10.1097/INF.0000000000002888. PMID: 32925547.

5) Lopez Bernal J, Andrews N, Gower C, Robertson C, Stowe J, Tessier E, Simmons R, Cottrell S, Roberts R, O’Doherty M, Brown K, Cameron C, Stockton D, McMenamin J, Ramsay M. Effectiveness of the Pfizer-BioNTech and Oxford-AstraZeneca vaccines on covid-19 related symptoms, hospital admissions, and mortality in older adults in England: test negative case-control study. BMJ. 2021 May 13;373:n1088. doi: 10.1136/bmj.n1088. PMID: 33985964; PMCID: PMC8116636.

6) Haas EJ, Angulo FJ, McLaughlin JM, Anis E, Singer SR, Khan F, Brooks N, Smaja M, Mircus G, Pan K, Southern J, Swerdlow DL, Jodar L, Levy Y, Alroy-Preis S. Impact and effectiveness of mRNA BNT162b2 vaccine against SARS-CoV-2 infections and COVID-19 cases, hospitalisations, and deaths following a nationwide vaccination campaign in Israel: an observational study using national surveillance data. Lancet. 2021 May 15;397(10287):1819–1829. doi: 10.1016/S0140-6736(21)00947-8. Epub 2021 May 5. Erratum in: Lancet. 2021 Jul 17;398(10296):212. PMID: 33964222; PMCID: PMC8099315.

7) Painel Rio COVID-19. Available at : https://experience.arcgis.com/experience/38efc69787a346959c931568bd9e2cc4 Accessed on September 03, 2021.

8) WHO Coronavirus (COVID-19) Dashboard. Available at: https://covid19.who.int/ Accessed on September 2, 2021.

9) Hodgson SH, Mansatta K, Mallett G, Harris V, Emary KRW, Pollard AJ. What defines an efficacious COVID-19 vaccine? A review of the challenges assessing the clinical efficacy of vaccines against SARS-CoV-2. Lancet Infect Dis. 2021 Feb;21(2):e26–e35. doi: 10.1016/S1473-3099(20)30773-8. Epub 2020 Oct 27. PMID: 33125914; PMCID: PMC7837315.

10) Hall VJ, Foulkes S, Saei A, Andrews N, Oguti B, Charlett A, Wellington E, Stowe J, Gillson N, Atti A, Islam J, Karagiannis I, Munro K, Khawam J, Chand MA, Brown CS, Ramsay M, Lopez-Bernal J, Hopkins S; SIREN Study Group. COVID-19 vaccine coverage in health-care workers in England and effectiveness of BNT162b2 mRNA vaccine against infection (SIREN): a prospective, multicentre, cohort study. Lancet. 2021 May 8;397(10286):1725–1735. doi: 10.1016/S0140-6736(21)00790-X. Epub 2021 Apr 23. PMID: 33901423; PMCID: PMC8064668.

